# DIP: Natural History Model for Major Depression with Incidence and Prevalence

**DOI:** 10.1101/2021.03.11.21253279

**Authors:** Melike Yildirim, Bradley N Gaynes, Pinar Keskinocak, Brian W Pence, Julie Swann

**Affiliations:** School of Industrial and Systems Engineering and Center for Health and Humanitarian Systems, Georgia Institute of Technology, Atlanta, Georgia, USA; Department of Psychiatry, University of North Carolina at Chapel Hill, Chapel Hill, North Carolina, USA; Department of Environmental Health, Rollins School of Public Health, Emory University, Atlanta, Georgia, USA; Gillings School of Global Public Health, University of North Carolina at Chapel Hill, Chapel Hill, North Carolina, USA; Department of Industrial and Systems Engineering, North Carolina State University, Raleigh, North Carolina, USA

**Keywords:** Major depression, Natural history, Prevalence, Incidence, Recall bias, Lifetime prevalence

## Abstract

**Background:** Major depression is a treatable disease, and untreated depression can lead to serious health complications. Therefore, prevention, early identification, and treatment efforts are essential. Natural history models can be utilized to make informed decisions about interventions and treatments of major depression.

**Methods:** We propose a natural history model of major depression. We use steady-state analysis to study the discrete-time Markov chain model. For this purpose, we solved differential equations and tested the parameter and transition probabilities empirically.

**Results:** We showed that bias in parameters might collectively cause a significant mismatch in a model. If incidence is correct, then lifetime prevalence is 33.2% for females and 20.5% for males, which is higher than reported values. If prevalence is correct, then incidence is .0008 for females and .00065 for males, which is lower than reported values. The model can achieve feasibility if incidence is at low levels and recall bias of the lifetime prevalence is quantified to be 31.9% for females and 16.3% for males.

**Limitations:** Model is limited to major depression, and patients who have other types of depression are assumed healthy. We assume that transition probabilities (except incidence rates) are correct.

**Conclusion:** We constructed a preliminary model for the natural history of major depression. We determine the lifetime prevalence are underestimated. We conclude that the average incidence rates may be underestimated for males. Our findings mathematically prove the arguments around the potential discordance between reported incidence and lifetime prevalence rates.

## Introduction

Major depression is a common mental illness, which affects roughly 17.3 million adults in the United States. It is more prevalent among women (10.2%) than men (6.2%) (Kessler et al., 2010). Major depression (or simply, depression) is also a leading cause of disability (Kessler et al., 1999). The annual direct and indirect costs of depression, which are mostly caused by decreased productivity and increased healthcare utilization, were estimated at $210.5 billion in 2010 (Greenberg et al., 2015).

Although major depression is a widespread disease, only 33% to 50% (Harman et al., 2006; Kessler et al., 2003; Pincus et al., 1998) of patients are diagnosed or receive adequate treatment in primary care settings. If it is not detected and treated, it can cause functional impairment and contribute to poor health outcomes. Therefore, prevention and treatment efforts are essential. Natural history models, which describe how diseases develop and progress over time, can help inform decisions about interventions and treatments. A well-established natural history model can be a powerful tool to project the disease burden and evaluate the effectiveness of intervention strategies. Projection of the potential benefit of intervention programs provides insight into the pros and cons of the promotion of such efforts. Information obtained from the natural history model can play an important role in answering policy questions that are not easily obtained from randomized control trials or clinical observational studies.

The natural history model for major depression has been developed for different study cohorts (e.g., 40-year-old primary care patients (Valenstein et al., 2001)) in the literature. However, obtaining a rigorous natural history model is challenging because of the disparities between published data. Annual incidence estimates are between 2.3 to 15.9 per 1000 people for major depression (Eaton et al., 1997; Mattisson et al., 2005; Murphy et al., 2002), and lifetime prevalence is usually reported as ranging from 10% to 20% (Bland, 1992; Kessler et al., 2003; Kessler et al., 2010). However, It is reported (Eaton et al., 1997) that incidence rates from the Baltimore Epidemiologic Catchment Area (ECA) (which are lower than the estimates from National Comorbidity Survey Replication (NCS-R)) would result in a 50% lifetime prevalence, which is higher than reported in other literature (Kessler et al., 2003). This infeasibility is consistent with other researchers, who have also identified a mismatch between reported incidence and lifetime prevalence (Kessler et al., 2003; Kessler et al., 2010; Patten, 2009; Takayanagi et al., 2014). Also, the 12-month prevalence of major depression bias was predicted as up to 20% (Patten et al., 2012), which is inconsistent with lifetime prevalence values. A systematic investigation is essential to clarify the inconsistencies due to misestimation of parameters.

One potential cause of the mismatch is recall bias on episodes of depression (Patten et al., 2012; Takayanagi et al., 2014; Wells and Horwood, 2004). Community-based surveys are commonly used to quantify the lifetime prevalence of major depression. Estimates about the lifetime prevalence are obtained from survey participants who recall any episode of disease over their entire lifetime. In the literature, the recall bias of lifetime prevalence of major depression was estimated as 35-291% (Andrews et al., 1999; Foley et al., 1998; Giuffra and Risch, 1994; Knauper and Wittchen, 1994; Kruijshaar et al., 2005; Patten, 2003, 2009; Takayanagi et al., 2014).

We build a revised a natural history model of major depression with incidence and prevalence (DIP) that mimics the disease progression without interventions. We utilize a Markov chain model with model inputs derived from published data, and we mathematically show how parameters can be mismatched and calibrated. Our study extends prior models (Valenstein et al., 2001) by performing a systematic investigation of parameters and calibrations to adapt the model to the current US adult population. We deliver a natural history model with calibrated parameters that are feasible as a system.

## Methods

### Natural History Model

We propose a natural history model of major depression with incidence and prevalence (DIP), which consists of four health states: healthy, depression, remission, and death (Figure 1). The healthy state is defined for people who have never had a major depressive disorder in their lifetime. The remission state contains patients with a history of depression who have not satisfied the diagnostic criteria for major depression in the past 12 months. Patients who fully recover from depression do not transition back to the healthy state. The death state is defined for all death causes, including depression-related and not depression-related. From each state, there exists a transition to the death state. Lifetime prevalence is the sum of the proportion of the population in the depression and remission states.

**Figure 1.**
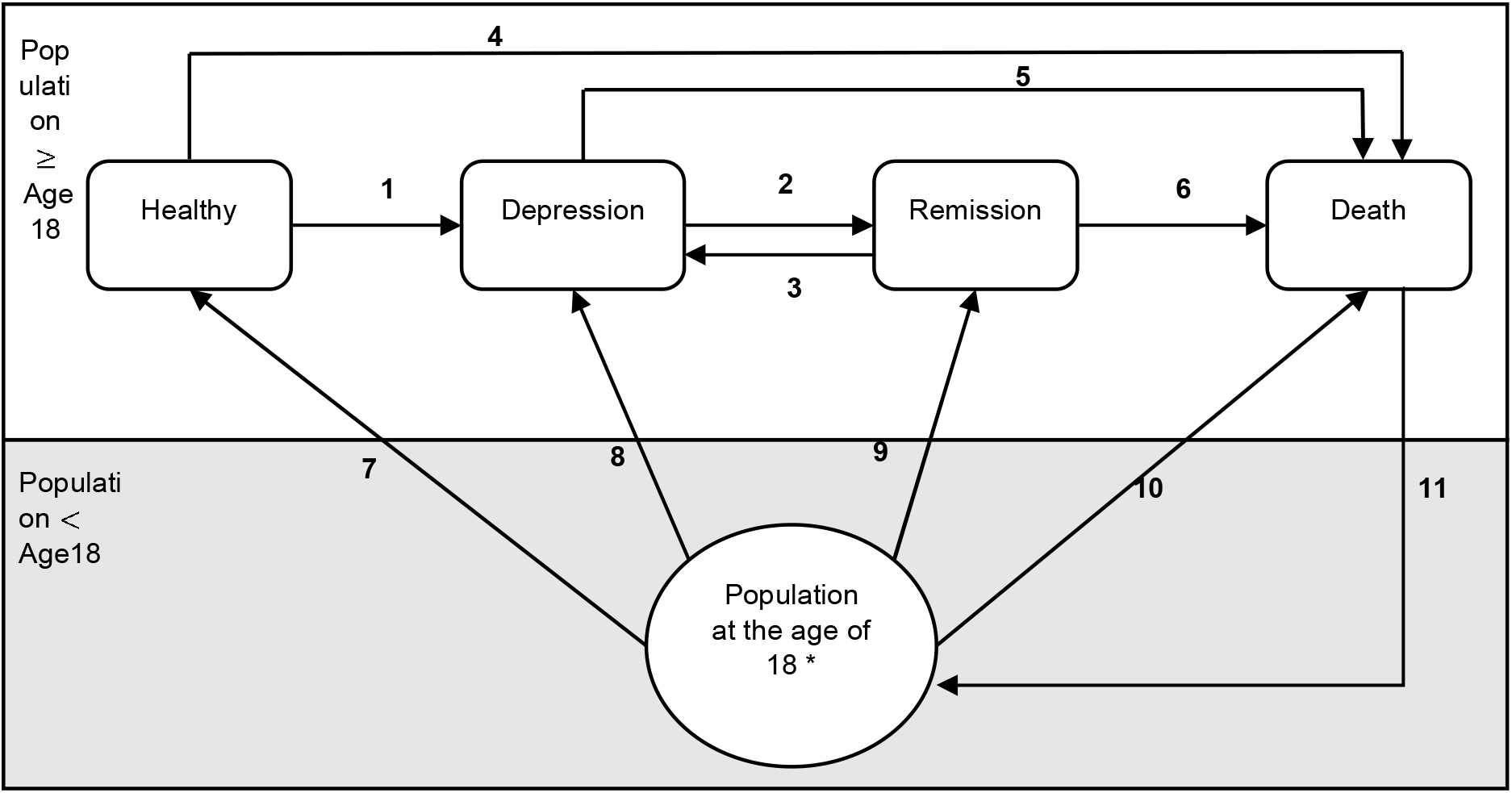
Depiction of the Markov model for the natural history of major depression. Notes: Boxes represent health states; arrows represent allowed transitions between states * This is a dummy state.

We utilize a discrete-time Markov chain model with a sequence of stochastic and state-to-state transitions. Patients are allowed to transition between health states at the end of each year. Our study population consists of adults age 18 or older, which is consistent with published data (e.g., prevalence, incidence) in the literature.

We use a constant size population and allow a new individual to enter the system when a person died (Bush and Zaremba, 1971). A dummy state is created to keep the population constant in every Markov cycle. Individuals enter into the system based on the average mortality rate of the adult population in the US. The newly introduced population is assigned initial states based on prevalence (arcs 7-10 in Figure 1) and enters the system at age 18.

### Model Parameterization

We initially derive the model parameters from highly cited studies including the nationally representative samples of US cohort studies of the NCS-R (Kessler et al., 2010) and ECA (Eaton et al., 2007). The rates are generally consistent with others; for example, the prevalences of depression from NCS-R are similar to the reported percentages from the National Institute of Mental Health. We use annual transition probabilities specific to males and females throughout the model, except for remission (Table 1), which has identical values for both genders.

**Table 1.** Transition probabilities of major depression: Markov model.

| Definition | Transitions | Parameter mean<br>[Female, Male] | Reference |
| --- | --- | --- | --- |
| Incidence | 1 | [0.0039, 0.0021] | ((Eaton et al., 2007) |
| Achieving remission | 2 | 0.45 | ((Brodaty et al., 1993;<br>Thase et al., 2005;<br>Whiteford et al., 2013)<sup>(a - b)</sup> |
| Recurrence of depression | 3 | [0.324, 0.281] | (Kessler et al., 1994)<sup>c</sup> |
| Mortality for general<br>population<br>(healthy or in remission) | 4, 6 | [0.016, 0.017] | (Arias et al., 2017)<sup>d</sup> |
| Mortality for depressive<br>patients | 5 | [0.025, 0.0269] | (Arias et al., 2017; Cuijpers<br>et al., 2014)<sup>(e, f)</sup> |
| Prevalence at the age of 18<br>(Healthy, Dep., Rem., Death) | 7, 8, 9, 10 | [(0.763, 0.136,<br>0.101, 0.000305),<br>(0.848, 0.072,<br>0.079, 0.000751)] | (Arias et al., 2017; Kessler<br>et al., 2010) |
| Reentrance (refer text for<br>more information) | 11 | 1 |  |

<sup>a</sup> Full remission rates for untreated patients are 0.37 (Whiteford et al., 2013) and for treated patients 0.47 (Thase et al., 2005). It is assumed that 65% of the patients are diagnosed (Simon and VonKorff, 1995) and, one-third of the diagnosed patients received treatment (Waitzfelder et al., 2018). The full remission rate is, on average, 0.39.
<sup>b</sup> Partial remission with the existence of residual symptoms is seen for 24% of the population in 3.8 years follow-up (Brodaty et al., 1993), which corresponds to the rate of 6.32% in a year.
<sup>c</sup> Probability for the population in the age range 15-54.
<sup>d</sup> Weighted average.
<sup>e</sup> Denotes that this source is a primary source.
<sup>f</sup> Denotes that this source used secondarily for an adjustment (multiplier) to the primary source.

We compute the weighted average of the age-specific death rate (Arias et al., 2017) based on the proportion of each age in the standard US population. Depressive patients are more likely to have any other comorbid condition; accordingly, they have an elevated risk of death compared to healthy patients. Therefore, their lifetime may be shorter than patients without depression. In our analysis, we use the relative risk of mortality in depressed patients as 1.58 compared to the non-depressed population (Cuijpers et al., 2014).

In the main results, we focus on the general steady state, that is, where the system would be stable. Note that for a given initial prevalence, the feasible pairs of incidence and lifetime prevalence in steady state can be calculated. In supplemental analysis, we parameterize the model with age-specific rates for incidence, lifetime prevalence, or recall bias (see the Supplement).

### Validation

We use a stationary Markov model, which implies that the transition probabilities are identical for every Markov cycle. A unique equilibrium exists regardless of initial distribution as a result of the features of the Markov model (irreducible and aperiodic; refer to Ross (Ross, 2010) for additional information). The equilibrium represents the fixed distribution of the population that is eventually observed. Steady-state analysis is used to obtain that equilibrium, and this can be used to test parameters and transition probabilities empirically. The result of the steady-state analysis is the distribution of people in each state of the system, which corresponds to prevalence. For the age-specific model, we used transient analysis to determine the state of the system after a set of years.

To examine the model with parameters obtained from the literature, we build two sets of linear equations, with one for each gender (Appendix 1 in Supplementary file) for the Markov model. We conduct our steady-state calculations using R (version 3.6.2) (Team, 2013). The equations are infeasible with observed prevalence (steady-state distribution) data which is P_female_ (healthy, depression, remission, death) = (0.755, 0.102, 0.127, 0.016) and P_male_ (healthy, depression, remission, death) = (0.832, 0.062, 0.089, 0.017) (Arias et al., 2017; Kessler et al., 2010; Kessler et al., 1997). Therefore, we further calibrate the parameters to build a model.

### Calibration

As documented (Eaton et al., 1997), an infeasible system of equations for depression can be caused by different reasons, e.g., the prevalence rate is too high, the incidence is too low, or both. In the following sections, we hypothesize and evaluate each of these cases.

In hypothesis 1, we assume that the observed lifetime prevalence of each state obtained from the literature is correct along with the initial prevalence at age 18. Therefore, one or more biased transition probabilities for incidence may lead to inconsistency between parameters. Thus, we calibrate incidence rates and determine the steady-state distribution of the Markov model for average values, comparing the results to the literature.

In hypothesis 2, we assume that reported transition probabilities (including for incidence) are correct, as is the initial prevalence at the age of 18 (Kessler et al., 2010; Kruijshaar et al., 2005). We focus on finding fitted values for prevalence for each model state and compare to the evidence from the literature. We calculate the steady-state distribution of the Markov model with average values.

In hypothesis 3, because of the high discordance between calculated and reported rates, we evaluate the case that both incidence and lifetime prevalence are incorrect. We perform the calibration by examining scenarios where the incidence is equal to the mean, lower, and upper bound obtained from the Baltimore ECA study (Eaton et al., 2007). The steady-state distribution is recalculated for the range of incidence values. We assume that recall bias may exist, and we calculate the amount needed for prevalence (annual or lifetime) values to fit within bounds.

## Results

### Hypothesis 1: Prevalence rates are correct

Assuming the stated prevalences in each state, we calculate the annual incidence rates from the steady-state distribution in the model as .0008 and .00065 for females, and males, respectively. Compared to the reported average incidence rates (Table 1) of .0039 and .0021, the incidence rates from the model calculations were lower (∼79% lower for females and ∼69% for males). This unexpected gap also exists between the calculated incidence and the stated lower bound from the 95% confidence interval of the incidence rates in the Baltimore ECA study (the lower bounds are .0029 and .0013, for females and males, respectively) (Eaton et al., 2007).

The steady-state equations yield all feasible pairs of incidence and lifetime prevalence, given initial conditions, recurrence, remission, and mortality. Figure 2 displays all feasible pairs, indicating the incidence and lifetime prevalence under Hypothesis 1.

**Figure 2.**
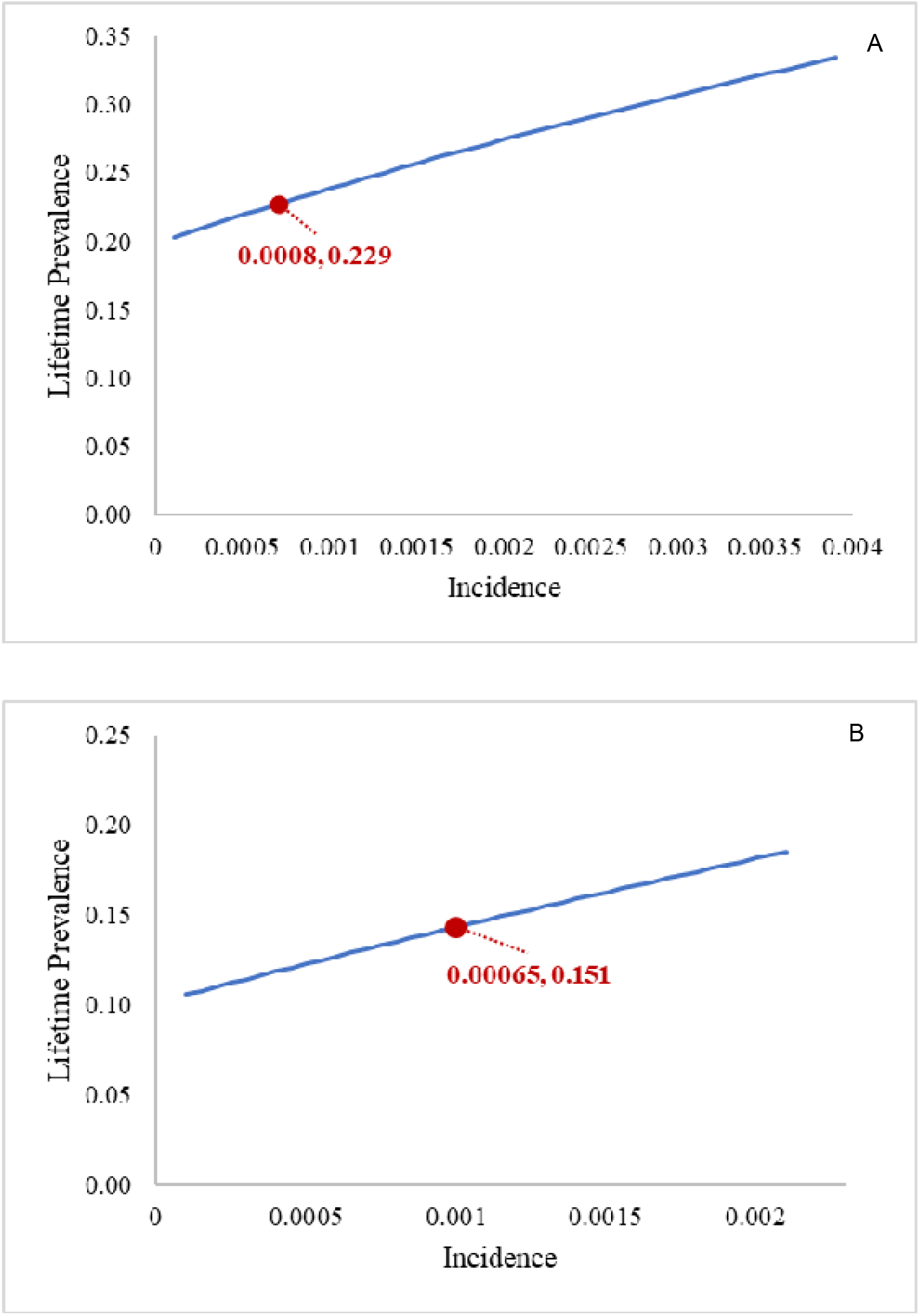
Feasible pairs of incidence and lifetime prevalence, with initial conditions, recurrence, remission, and mortality. Note: Labeled pair shows values resulting from prevalence rates from the literature and calculated incidence; A shows Female, and B shows Male.

### Hypothesis 2: Transition probabilities for incidence are correct

Given incidence rates, we calculate the lifetime prevalence of depression (depression plus remission) from the model as 33.2% for females and 20.5% for males. However, the observed prevalence from national surveys was 22.9% and 15.1%, respectively (Kessler et al., 2010). The calculated lifetime prevalence rates with known incidence are 45% higher for females and 35.9% higher for males than reported values.

Using the Markov model and the transition probabilities, we plotted the prevalence to compare the expected values with the observed values. Figure 3 shows that the transition probabilities from the literature lead to higher depression and remission prevalence than is stated in the literature. Additionally, we observed that fewer individuals had never experienced an episode of depression in their lifetime (65.1% calculated vs. 75.5% observed for females and 77.7% calculated vs. 83.2% observed for males) in the model results compared to the reported values.

**Figure 3.**
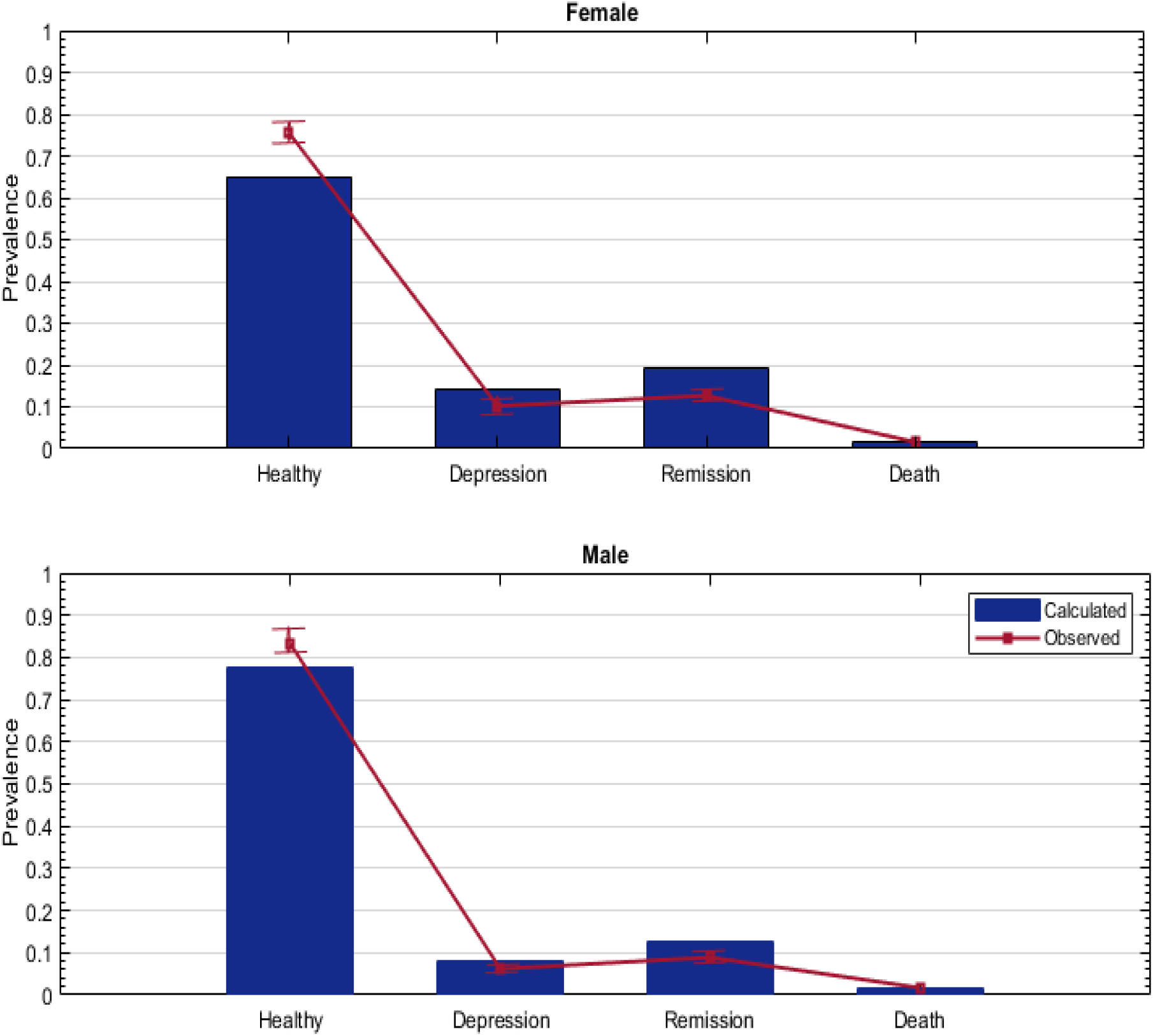
Comparison of calculated prevalence from the Markov model and observed prevalence from the literature for each health state assuming that incidence is correct, where the observed proportion of healthy people is higher than the steady state analysis. * Observed prevalence rates are obtained from Kessler et al. (1) and calculated as 95% CI.

### Hypothesis 3: Prevalence and incidence rates are biased

Using several scenarios for incidence values, we obtain calculated values for prevalence from the steady-state model and recall bias necessary to match reported prevalence values.

We quantified that lifetime prevalence in steady state ranged from 30.2% to 36.4% for females (vs. 17.6 to 24.5% for males) and 12-month prevalence ranged from 12.8% to 15.4% for females (vs. 6.8 to 9.6% for male). Even with a low incidence rate, the lifetime prevalence values in steady state are higher (as shown in Figure S1) than the values in the literature, which are 22.9% and 15.1%, respectively (Kessler et al., 2010), for females and males.

We calculated the recall bias of the lifetime prevalence based on the gap between the estimated lifetime prevalence from the Markov model (as shown in Table 2) and the observed lifetime prevalence in the literature.

**Table 2.** Calibration of the Markov model and calculated prevalence and recall bias

| **Gender** | **Low** | **Medium** | **High** |
| --- | --- | --- | --- |
|  | **Incidence\*** |  |  |
| Female | 0.0029 | 0.0039 | 0.0051 |
| Male | 0.0013 | 0.0021 | 0.0033 |
| **Calculations from Markov Model** |  |  |  |
|  | **Lifetime Prevalence (%)** |  |  |
| Female | 30.2 | 33.2 | 36.4 |
| Male | 17.6 | 20.5 | 24.5 |
|  | **12-month Prevalence (%)** |  |  |
| Female | 12.8 | 14.1 | 15.4 |
| Male | 6.8 | 8.0 | 9.6 |
| **Calculated Recall Bias** |  |  |  |
|  | **Lifetime Prevalence (%)** |  |  |
| Female | 31.9 | 45.0 | 59.0 |
| Male | 16.3 | 35.8 | 62.3 |
|  | **12-month Prevalence (%)** |  |  |
| Female | 25.5 | 38.2 | 51.0 |
| Male | 9.0 | 29.0 | 54.8 |

\* 95% CI of the incidence rate in the Baltimore ECA study (Eaton et al., 2007)

The gap between calculated and observed lifetime prevalence ranged from 31.9% to 59% for females and 16.3 to 62.3% for males. Additionally, for the lower bound of incidence (which was 25.6% and 38.1% lower than the average incidence, respectively for females and males), we calculated the recall bias for the 12-month prevalence of major depression as 25.5% for females and 9% for males. We also had age-specific analysis, including additional parameter settings of higher mortality for people who are depressed or with increased incidence for the younger population.

## Discussion

In hypothesis 1, we found that assuming prevalence values from the literature are correct resulted in an incidence rate that is lower than observed in the literature (Eaton et al., 2007), so this hypothesis seems unlikely to be true. In hypothesis 2, we found that assuming incidence values from the literature are correct resulted in lifetime prevalence values that are higher than are observed in the literature (Kessler et al., 2010), so this hypothesis also seems unlikely to be true. In hypothesis 3, we reported recall bias estimates for extreme values of incidence rates. Even with the lower bound of incidence, we find that the lifetime prevalence in steady state is higher than that observed in practice. This suggests that the lifetime prevalence that is reported in practice may be underestimated. We find that the model can be in steady state with extreme values of incidence and with recall bias that increases with age.

One of the possible explanations of high prevalence among young population is the “cohort effect”. New generations may have an elevated risk of depression. It is (Wickramaratne et al., 1989) showed an increased number of cases in the birth cohort born during the years 1935-1945. However, if such an increasing trend for incidence exists, we should see this rising trend move to another subsequent age group over the years, and it is not observed in 2005 for adults over 30 (Twenge et al., 2019).

On the other hand, we may see fewer patients with depression in older age groups because of the elevated risk of mortality of depressive patients. We account for increased mortality in patients with depression in our model, and we conclude this alone does not explain the results. Our age-specific analysis further supports this conclusion.

Cohort effects and increased mortality of depressive patients are not enough to explain the gap between observed and calculated lifetime prevalence from reported rates (Appendices 2 and 3) (Patten et al., 2010). There may be other factors that contribute to the pattern, such as the changes in diagnostic criteria. In the past half-century, problems of anxiety started to be called depression (Horwitz, 2010).

In our age-specific analysis, we showed that the recall bias increases until the age of 79 (Table S3). To be more conservative on the calculation of bias, we used prevalence from NCS-R (Kessler et al., 2010), which reported relatively higher rates than the study (ECA (Eaton et al., 2007)) that incidence rates are obtained. Furthermore, we observed that recall bias still exists even if the mortality risk of people with depression was much higher than others, e.g., a 4-fold increase, (Figures S2 and S3 in Supplementary) and if there is a significant (e.g., 6-fold) increase in incidence among younger birth cohorts (Figures S4 and S5 in Supplementary).

Recall problems may increase with age because of a high risk for cognitive decline. Mental health problems may also fade in the face of physical ailments that increase with age. As well as age, the number of previous episodes and time since the last episode may affect reporting. It is been observed that 10% of patients did not report their depression episodes at onset (Patten et al., 2012), and the recall bias can exist when the recall period is as short as one week (Zanni, 2007). In our results, we calculated relatively low recall bias rates for males would be necessary to match incidence and lifetime prevalence in the steady state. This does not seem true, because some studies show that females have a better memory than males (Lundervold et al., 2014). In the age-specific analysis, we observed that men have higher recall bias in older ages (≥79 years old) than females (Table S3 and Figure S5 in the Supplement). However, men may hide psychological problems, and they may be reluctant to seek help for their conditions (Lee and Owens, 2002; O’Brien et al., 2005). This indicates that the incidence rate may be underestimated for males. Additionally, another concern is measurement bias, which may lead to underestimating the disease burden among men. In general, tools and questions that are used in surveys detect the symptoms the same way for men and women (Smith et al., 2018).

One of the recent studies showed a relatively higher lifetime prevalence (14.7% for males and 26.1% for females) of major depression than previous studies, using The National Epidemiologic Survey on Alcohol and Related Conditions III (Hasin et al., 2018). However, concerns about the underreporting rates still exist even with cumulative estimates (Wells and Horwood, 2004). Cumulative evaluations from multiple interviews may also underestimate the true lifetime prevalence of major depression because of the patients who did not recall their lifetime event in all interviews (Takayanagi et al., 2014). Therefore, the lifetime prevalence values estimated from simulation studies were higher rates than the general population surveys, either one-time retrospective or cumulative evaluations. (Kruijshaar et al., 2005) estimated the lifetime prevalence of 20% for men and 30% for women from a microsimulation study.

Our model provides a preliminary framework for the natural history of major depression. Additionally, we quantified the calibration for incidence and lifetime prevalence. Our findings mathematically prove and support the arguments around the potential discordance between incidence and lifetime prevalence rates, which can be solved in part by adjusting for recall bias. Markov models can also be used by other researchers to estimate recall bias or other adjustments needed to ensure the feasibility of parameters for models. Furthermore, our models are available for others, and we provide steady-state calculations as an R script (Appendix 4 in Supplementary file).

### Limitations

This study has several limitations about the model structure, data inputs, and calibration process. First, we have included a relatively small number of states rather than characterizing based on the severity of depression (low, moderate, and high) and recovery procedure (partial remission, full remission). Additionally, our model is limited to major depression, and patients who have other types of depression are assumed healthy. The stationary (time-homogeneous) assumption of the model may not entirely match with practice; however, it is a useful to show the stability of disease distribution and quantify adjustments (Bush and Zaremba, 1971). Besides, we support the argument by providing age-specific analysis of lifetime prevalence with time-dependent parameters (e.g., incidence, prevalence, and mortality) in Appendix 2 in the Supplementary file. Furthermore, in all the steps of the calibration process, we assume that transition probabilities (except incidence rates) are correct, while we obtained the data from various studies from different years.

## Conclusions

Average incidence estimates have often seemed unrealistically high (Eaton et al., 1997; Patten, 2008) because they would lead to excessively high lifetime prevalence (33.2% for females and 20.5% for males); however, lifetime prevalence is probably much higher (Kruijshaar et al., 2005; Takayanagi et al., 2014) than reported in general population surveys.

In the literature, studies were reported of recall bias of 38% or more (Andrews et al., 1999; Kruijshaar et al., 2005). Our rates are consistent with this, while additionally suggesting that the average incidence rates may be underestimated for males.

Because reported lifetime prevalence is an underestimate, this causes an overestimation of the healthy population. Optimistic assumptions about the prevalence of major depression have significant consequences for underestimating the burden of disease and the benefit of preventive care. The high prevalence of major depression is a public health concern, and its burden higher than expected. Our model could be utilized to analyze the natural major depression behavior, treatment decisions, or estimate the costs associated with screening.

## Data Availability

All data used in the study is publicly available as described in the paper.

## Acknowledgements

The authors are grateful to the Editors and anonymous reviewers for their thoughtful, constructive comments. We gratefully acknowledge Hannah Smalley, Ph.D., for her comments and suggestions on the first draft of this manuscript.

