## Supplementary file for "DIP: Natural History Model for Major Depression with Incidence and Prevalence"

This supplementary material file has been created by the authors to give readers additional information about the research article with the above title.

**Appendix 1.** System of linear equations

**Figure S1.** Calculated lifetime prevalence versus observed lifetime prevalence without recall bias

**Appendix 2.** Age-specific analysis of lifetime prevalence

**Table S1.** Calculated lifetime prevalence

**Table S2.** Transition probabilities of age-specific Markov model

**Table S3.** Calculated recall biases

**Figure S2.** Lifetime prevalence for female

**Figure S3.** Lifetime prevalence for male

**Appendix 3.** Analysis with cohort effects

**Table S4.** Modified incidence values used in consideration of cohort effects from,(Eaton et al., 2007; Twenge et al., 2019; Wickramaratne et al., 1989) with calculated and observed values for lifetime prevalence

**Table S5.** Modified incidence under the consideration of cohort effects from, (Patten et al., 2010; Twenge et al., 2019; Wickramaratne et al., 1989) with calculated and observed values for lifetime prevalence

**Table S6.** Calculated recall biases with cohort effects

**Figure S4.** Lifetime prevalence in 2001 for modified incidence rates with cohort effect and relative mortality rate (Female)

**Figure S5.** Lifetime prevalence in 2001 for modified incidence rates with cohort effect and relative mortality rate (Male)

**Appendix 4.** R script

**References**

**Appendix 1.** System of linear equations

**Female (1a-1d***)*

$0.980 P_{healthy}+0.763 P_{death}=P_{healthy}$

$0.017 P_{healthy}+0.0269 P_{depression}+0.017 P_{remission}+0.000751 P_{death} =P_{death}$

$0.46 P_{depression}+0.702 P_{remission}+0.079P_{death}=P_{remission}$

$0.0021 P_{healthy}+0.513 P_{depression}+0.281 P_{remission}+0.072 P_{death} =P_{depression}$

$0.9809 P_{healthy}+0.848 P_{death}=P_{healthy}$

$0.46 P_{depression}+0.66 P_{remission}+0.101P_{death}=P_{remission}$

$0.016 P_{healthy}+0.025 P_{depression}+0.016 P_{remission}+0.000305 P_{death} =P_{death}$

$0.0039 P_{healthy}+0.515 P_{depression}+0.324 P_{remission}+0.136 P_{death} =P_{depression}$

**Male (2a-2d)**

Equations are infeasible where the steady state distribution (P_healthy_, P_depression_, P_remission_, P_death_) = (0.755, 0.102, 0.127, 0.016) and where (P_healthy_, P_depression_, P_remission_, P_death_) = (0.832, 0.062, 0.089, 0.017), respectively, for the female and male populations (Arias et al., 2017; Cuijpers et al., 2014; Kessler et al., 2010).

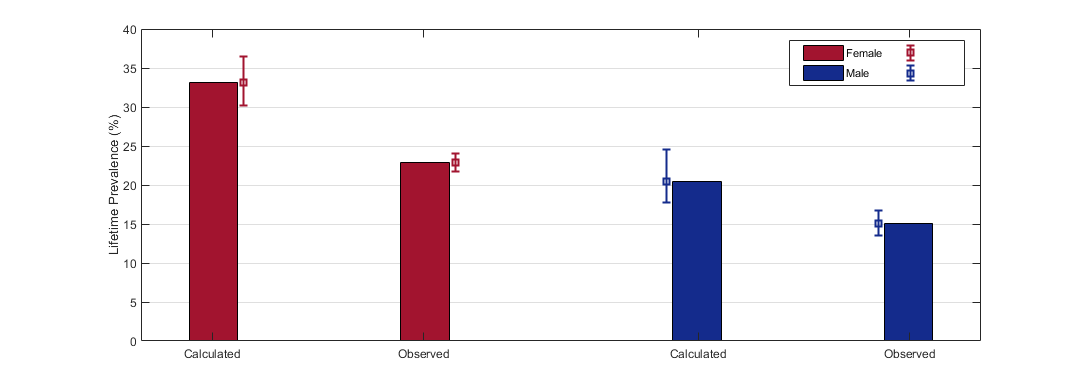
**Figure S1.** Calculated lifetime prevalence versus observed lifetime prevalence without recall bias.

*** Observed lifetime prevalence rates are obtained from Kessler et al. (Kessler et al., 2010) and calculated as 95% CI; calculated lifetime prevalence and the range are obtained using varying incidence rates as shown in manuscript Table 2.**

**Appendix 2. Age-specific analysis of lifetime prevalence**

We performed age-specific analysis for the natural history model from Figure 1. We updated some of the parameters (e.g., incidence, prevalence, and mortality), as shown in Table S1.

We would like to see the distribution within states after a period of time; thus, we performed transient analysis capturing the transitions for the specific number of years. We calculated lifetime prevalence up to age X for values of 20, 34, 49, 64, 79, and 90. Lifetime prevalence at age X indicates the proportion of a population that met the criteria for Major Depression at least once during the time between ages 18 and X.

In the overall study, we used the average mortality risk ratio as 1.58 for depressed patients (Cuijpers et al., 2014). In this part of the study, we extend our analysis with mortality risk ratio up to 4.

**Table S1.** Transition probabilities of age-specific Markov model

|  |  | **Parameter (mean)** | | | | | | | |  |
| --- | --- | --- | --- | --- | --- | --- | --- | --- | --- | --- |
| **Definition** | **Transitions** | **Female** | | | | **Male** | | | | **Reference** |
|  |  | **18-29** | **30-44** | **45-64** | **65+** | **18-29** | **30-44** | **45-64** | **65+** |  |
| Incidence (95% CI) | 1 | 0.0045 (0.0019-0.0088) | 0.0065 (0.0045-0.0092) | 0.0024 (0.0011-0.0046) | 0.001 (0.0002-0.0028) | 0.0015 (0.0002 - 0.0052) | 0.0038 (0.002-0.0065) | 0.0009 (0.001-0.0032) | 0.0007 (0.00004-0.0037) | (Eaton et al., 2007) |
| Achieving remission | 2 | 0.45 | | | | | | | | (Brodaty et al., 1993; Thase et al., 2005; Whiteford et al., 2013) |
| Recurrence of depression | 3 | 0.324 | | | | 0.281 | | | | (Kessler et al., 1994) |
|  | | **18-34** | **35-49** | **50-64** | **65+** | **18-34** | **35-49** | **50-64** | **65+** |  |
| Mortality for general population | 4, 6 | 0.00054 | 0.00156 | 0.00527 | 0.0458 | 0.00131 | 0.002499 | 0.00865 | 0.052 | (Arias et al., 2017) |
| Mortality for depressive patients | 5 | 0.0008532 | 0.002465 | 0.00833 | 0.072 | 0.00207 | 0.00394 | 0.00137 | 0.0822 | (Arias et al., 2017; Cuijpers et al., 2014) |
| ^Ψ^Prevalence between the ages of 18-34*, 18-49**, 18-64***, 18-79**** | 7, 8, 9, 10 | *(0.763, 0.136, 0.101, 0.000305) | **(0.746, 0.124, 0.128, 0.00164) | ***(0.748, 0.114, 0.137, 0.00206) | ****(0.764, 0.102, 0.130, 0.00316) | *(0.848, 0.072, 0.079, 0.000751) | **(0.830, 0.073, 0.096, 0.00123) | ***(0.834, 0.068, 0.096, 0.00237) | ****(0.850, 0.060, 0.087, 0.00393) | (Arias et al., 2017; Eaton et al., 2007)^a^ |
| Reentrance (refer text for more information) | 11 | 1 | | | | | | | |  |

^Ψ^ 4 x 2 array of number refer to the 4 states (healthy, depression, remission, death).

^a^ Weighted average until ages 34, 49, 64, 79, population distribution obtained from (Lindsay M. Howden, May 2011)

CI – Confidence interval

In Table S2, we report the calculated lifetime prevalence values using mean incidence rates. Additionally, based on the upper and lower bounds (95% CI) of the incidence rate (Table S1), we calculate the error bars, as shown in Figures S2 and S3.

Figures S2 and S3, which also show the observed lifetime prevalence (Kessler et al., 2010), show that the calculated lifetime prevalence starts to drop after the age of 49, as the observed does. However, the difference between observed and calculated prevalence is increasing with age except for age 90.

Table S3 shows the recall bias (the difference between observed and calculated prevalence) that would be necessary for the system to be feasible, for low, medium, and high values of the incidence rate. For some ages (e.g., < 65), there are recall bias values that could be possible, especially for low and medium incidence. On the other hand, for the highest ages, the recall values above 100 are not possible, suggesting that recall bias alone does not explain the infeasibility in the system based on incidence, lifetime prevalence, and recall bias.

**Table S1.** Calculated Lifetime Prevalence (%) with

|  | Lifetime Prevalence^*^ | | Lifetime Prevalence^**^ | |
| --- | --- | --- | --- | --- |
| Age | Female | Male | Female | Male |
| 20 | 24.72 | 15.47 | 24.70 | 15.44 |
| 34 | 30.02 | 18.14 | 29.82 | 17.85 |
| 49 | 37.92 | 23.65 | 37.19 | 22.75 |
| 64 | 37.17 | 22.29 | 34.87 | 19.93 |
| 79 | 35.21 | 20.37 | 29.76 | 15.87 |
| 90 | 32.52 | 18.72 | 24.04 | 12.93 |

^*^Mortality risk ratio=1.58 and ^**^Mortality risk ratio=4.

**Figure S2.** Lifetime prevalence (%) for female
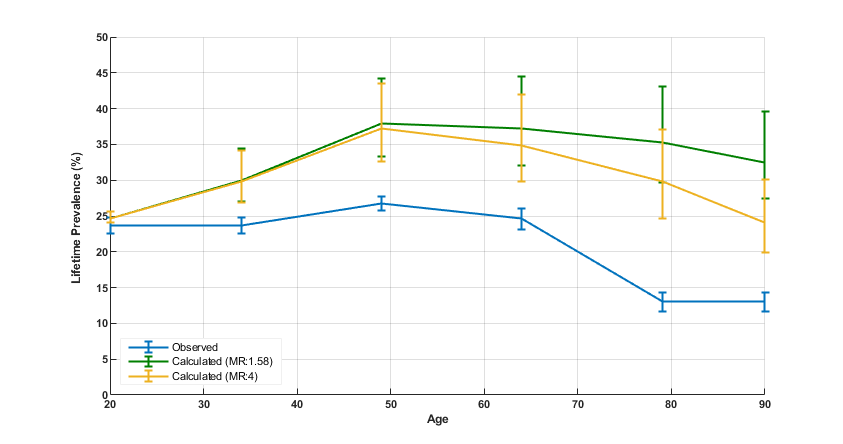

MR – Mortality Risk Ratio

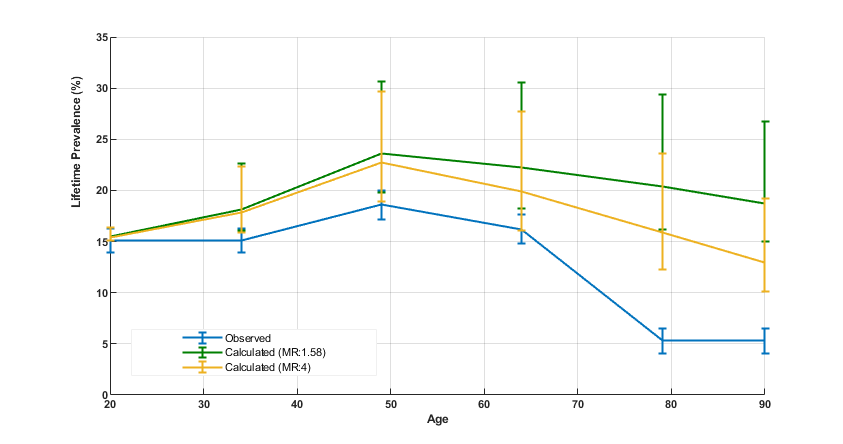
**Figure S3.** Lifetime prevalence (%) for male

MR – Mortality Risk Ratio

**Table S3.** Calculated Recall Biases

|  | **Female** | | | | | | **Male** | | | | | |
| --- | --- | --- | --- | --- | --- | --- | --- | --- | --- | --- | --- | --- |
|  | **Low Incidence** | | **Medium Incidence** | | **High Incidence** | | **Low Incidence** | | **Medium Incidence** | | **High Incidence** | |
| **Age** | *MR: 1.58* | *MR: 4* | *MR: 1.58* | *MR: 4* | *MR: 1.58* | *MR: 4* | *MR: 1.58* | *MR: 4* | *MR: 1.58* | *MR: 4* | *MR: 1.58* | *MR: 4* |
| **20** | 1.81 | 1.71 | 4.30 | 4.20 | 8.39 | 8.30 | 0.28 | 0.05 | 2.46 | 2.23 | 8.65 | 8.41 |
| **34** | 14.36 | 13.58 | 26.66 | 25.84 | 45.07 | 44.19 | 6.72 | 4.91 | 20.14 | 18.21 | 50.11 | 47.91 |
| **49** | 24.61 | 22.03 | 42.01 | 39.28 | 65.77 | 62.85 | 6.33 | 1.91 | 27.15 | 22.30 | 64.97 | 59.37 |
| **64** | 29.93 | 21.18 | 51.08 | 41.74 | 80.62 | 70.50 | 13.15 | 0.00 | 38.54 | 23.89 | 90.03 | 72.47 |
| **79** | 128.29 | 89.81 | 170.86 | 128.92 | 231.30 | 184.88 | 205.36 | 131.67 | 284.33 | 199.42 | 453.56 | 345.89 |
| **90** | 111.80 | 52.67 | 150.16 | 84.95 | 205.07 | 131.93 | 183.63 | 90.35 | 253.19 | 144.01 | 404.71 | 262.88 |

MR – Mortality Risk Ratio

**Appendix 3.** Analysis with cohort effects

Compared to previous cohorts, studies show (Twenge et al., 2019) that recent cohorts have had an increased incidence rate for the earlier ages, i.e., 18 to 29. On average, the increase corresponds to a 6% increase in annual incidence between generations in (Twenge et al., 2019). Other researchers have studied the increased incidence in later generations as a ratio of the earlier generation (Wickramaratne et al., 1989). In this analysis, we evaluated age cohort effects using both ways of measuring increased risk. For the analysis, we examine the state of the system in 2001.

We obtain the lifetime prevalence by year 2001 (Kessler et al., 2010) for birth cohorts born in the years 1920, 1931, 1946, 1960, or 1976. We calculated the incidence by age for specific birth cohorts from (Eaton et al., 2007; Twenge et al., 2019), where we specifically obtain the average incidence (by 6% increasing in subsequent years) in the early adult years (ages 18-29). After that age, we use the average incidence over the lifetime (Eaton et al., 2007). For each of the cohorts, we do a transient analysis from age 18 to their age in the year 2001, to calculate their lifetime prevalence until that point. For comparison, we also calculate the lifetime prevalence of depression without cohort effects for the same ages (using the same technique reported in Appendix 2 and obtained using incidence and prevalence values from Table S1).

In further analysis, we use incidence rates over a lifetime, as shown in Table S5. We use the birth cohorts from 1911 and 1922 as the baseline, and we compute the incidence for later birth cohorts according to the relative risk ratio obtained from (Patten et al., 2010). For example, the incidence for the cohort born in 1976 has an incidence rate 6.4 times that of the baseline generations.

|  | | Average Incidence | | | | Lifetime Prevalence | | | | | |
| --- | --- | --- | --- | --- | --- | --- | --- | --- | --- | --- | --- |
|  |  | In ages 18-29 | | In Lifetime (until 2001) | | Calculated with cohort effects ^++^ | | Calculated without cohort effects^++^ | | Observed | |
| Birth cohort | Age in 2001 | Female | Male | Female | Male | Female | Male | Female | Male | Female | Male |
| 1976 | 34 | 0.0041 | 0.0014 | 0.0049 | 0.0022 | 29.6% | 18.0% | 30.0% | 18.1% | 23.7% | 15.1% |
| 1960 | 50 | 0.0015 | 0.0005 | 0.0041 | 0.0025 | 34.9% | 22.8% | 37.9% | 23.7% | 26.7% | 18.6% |
| 1946 | 64 | 0.0007 | 0.0002 | 0.0034 | 0.0017 | 35.0% | 21.3% | 37.2% | 22.3% | 24.6% | 16.2% |
| 1931 | 79 | 0.0004 | 0.0001 | 0.0029 | 0.0015 | 32.5% | 19.3% | 35.2% | 20.4% | 13.0% | 5.3% |
| 1920 | 90 | 0.0002 | 0.0001 | 0.0019 | 0.0008 | 27.5% | 15.6% | 32.5% | 18.7% | 13.0% | 5.3% |

**Table S4.** Modified incidence values used in consideration of cohort effects from,(Eaton et al., 2007; Twenge et al., 2019; Wickramaratne et al., 1989) with calculated and observed values for lifetime prevalence.

^++^ Mortality Risk Ratio = 1.58.

**Table S5.** Modified incidence under the consideration of cohort effects from, (Patten et al., 2010; Twenge et al., 2019; Wickramaratne et al., 1989) with calculated and observed values for lifetime prevalence.

|  | |  | | Lifetime Prevalence | | | | | |
| --- | --- | --- | --- | --- | --- | --- | --- | --- | --- |
|  | | Average Incidence in Lifetime | | Calculated with cohort effects ^++^ | | Calculated without cohort effects^++^ | | Observed | |
| Birth Cohort | Relative Risk | Female | Male | Female | Male | Female | Male | Female | Male |
| 1967 | 6.4 | 0.00490 | 0.00216 | 29.6% | 18.1% | 26.1% | 15.9% | 23.7% | 15.1% |
| 1951 | 5.3 | 0.00400 | 0.00179 | 34.7% | 21.2% | 32.6% | 18.2% | 26.7% | 18.6% |
| 1936 | 1.7 | 0.00129 | 0.00057 | 28.6% | 17.6% | 37.0% | 23.2% | 24.6% | 16.2% |
| 1922 | Baseline | 0.00080 | 0.00034 | 24.7% | 15.0% | 37.6% | 22.6% | 13.0% | 5.3% |
| 1911 | Baseline | 0.00080 | 0.00034 | 23.6% | 14.1% | 35.5% | 20.5% | 13.0% | 5.3% |

^++^ Mortality Risk Ratio = 1.58.

**Table S6.** Calculated recall biases with cohort effects.

|  | Recall bias (%) | | | |
| --- | --- | --- | --- | --- |
|  | Calculated from Table C4 | | Calculated from Table C5 | |
| Birth cohort | Female | Male | Female | Male |
| 1976 | 24.9 | 19.2 |  | |
| 1967 |  | | 24.9 | 19.9 |
| 1960 | 30.7 | 22.6 |  | |
| 1951 |  | | 30.0 | 14.0 |
| 1946 | 42.3 | 31.5 |  | |
| 1936 |  | | 16.3 | 8.6 |
| 1931 | 150.0 | 264.2 |  | |
| 1922 |  | | 90.0 | 183.0 |
| 1920 | 111.5 | 194.3 |  | |
| 1911 |  | | 81.5 | 166.0 |

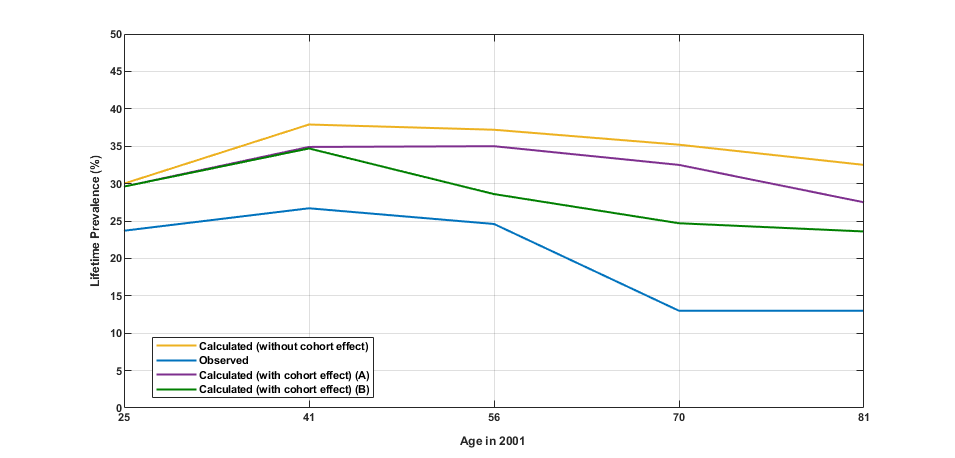
**Figure S4.** Lifetime prevalence in 2001 for modified incidence rates with cohort effect and relative mortality rate (Female).

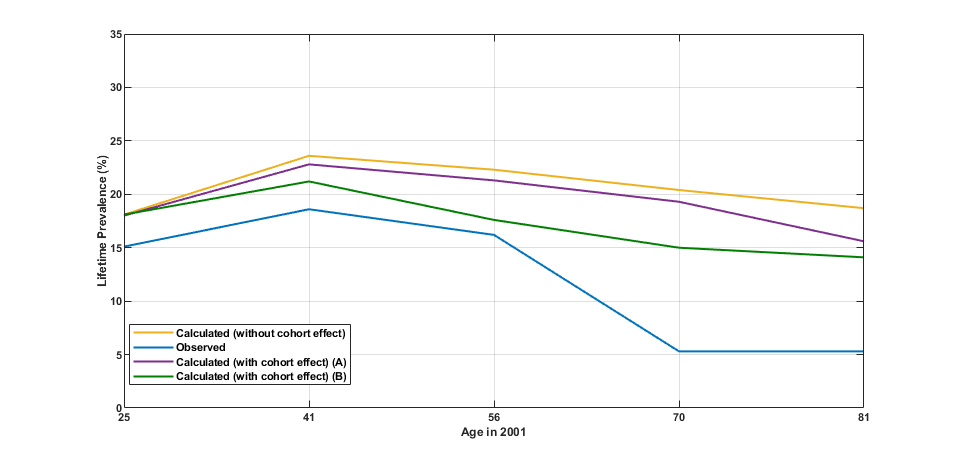
**Figure S5.** Lifetime prevalence in 2001 and recall bias for modified incidence rates with cohort effect and relative mortality rate (Male).

We illustrate the calculated lifetime prevalence with and without cohort effects up to year 2001 for each of several different age groups in Figures S4 (females) and S5 (males). The observed lifetime prevalence is provided in the figure as well. A key finding is that the recall bias (Table S6) is not higher than ~40% for cohorts born in 1936 or later, while for cohorts born earlier it would need to be unreasonably high (about 100% or more).

Thus, we conclude that birth cohorts and recall bias alone do not explain the gap between incidence and resulting lifetime prevalence for older generations. Other possibilities (that are outside the scope of this paper) include (1) that physical challenges overshadow the experience of or memory of mental health challenges so the mental health challenges are perceived as less important than they previously were; (2) different internal conceptualizations of depression for older generations, e.g., which may be related to language used in different time periods. We leave additional analysis to other researchers.

**Appendix 4.** R Script

#Table 1 (F-Female, M-Male)

incidence_F=0.0039

remission=0.45

recurrence_F=0.324

mortality_F=0.016

dep_mortality_F=0.025

pre_18_h_F=0.762695

pre_18_dep_F=0.136

pre_18_r_F=0.101

pre_18_d_F=0.000305

incidence_M=0.0021

recurrence_M=0.281

mortality_M=0.017

dep_mortality_M=0.0269

pre_18_h_M=0.848

pre_18_dep_M=0.072

pre_18_r_M=0.079

pre_18_d_M=0.000751

#__________________Hypothesis 1____________________

#Female

p = matrix(c((1-mortality_F-incidence_F),0,0,pre_18_h_F,

incidence_F,(1-remission-dep_mortality_F),recurrence_F,pre_18_dep_F,

0,remission,(1-recurrence_F-mortality_F),pre_18_r_F,

mortality_F,dep_mortality_F,mortality_F,pre_18_d_F),

nrow = 4,

ncol = 4)

n = ncol(p)

A = t(p - diag(n))

A = rbind(A, rep(1, n))

b = c(rep(0, n), 1)

mu = qr.solve(A, b)

print(mu)

#Male

p = matrix(c((1-mortality_M-incidence_M),0,0,pre_18_h_M,

incidence_M,(1-remission-dep_mortality_M),recurrence_M,pre_18_dep_M,

0,remission,(1-recurrence_M-mortality_M),pre_18_r_M,

mortality_M,dep_mortality_M,mortality_M,pre_18_d_M),

nrow = 4,

ncol = 4)

n = ncol(p)

A = t(p - diag(n))

A = rbind(A, rep(1, n))

b = c(rep(0, n), 1)

mu = qr.solve(A, b)

print(mu)

#__________________Hypothesis 2____________________

###Female

seq <- seq(0.0001,0.0039,by=0.0001)

for(incidence_F in seq)

{

p = matrix(c((1-mortality_F-incidence_F),0,0,pre_18_h_F,

incidence_F,(1-remission-dep_mortality_F),recurrence_F,pre_18_dep_F,

0,remission,(1-recurrence_F-mortality_F),pre_18_r_F,

mortality_F,dep_mortality_F,mortality_F,pre_18_d_F),

nrow = 4,

ncol = 4)

n = ncol(p)

A = t(p - diag(n))

A = rbind(A, rep(1, n))

b = c(rep(0, n), 1)

mu = qr.solve(A, b)

print(mu)

}

###Male

seq <- seq(0.0001,0.0021,by=0.00005)

for(incidence_M in seq)

{

p = matrix(c((1-mortality_M-incidence_M),0,0,pre_18_h_M,

incidence_M,(1-remission-dep_mortality_M),recurrence_M,pre_18_dep_M,

0,remission,(1-recurrence_M-mortality_M),pre_18_r_M,

mortality_M,dep_mortality_M,mortality_M,pre_18_d_M),

nrow = 4,

ncol = 4)

n = ncol(p)

A = t(p - diag(n))

A = rbind(A, rep(1, n))

b = c(rep(0, n), 1)

mu = qr.solve(A, b)

print(mu)

}

#__________________Hypothesis 3____________________

###Female

incidence_F_low=0.0029

incidence_F_high=0.0051

observed_lifetime_prevalence_F <- 0.229

p = matrix(c((1-mortality_F-incidence_F_low),0,0,pre_18_h_F,

incidence_F_low,(1-remission-dep_mortality_F),recurrence_F,pre_18_dep_F,

0,remission,(1-recurrence_F-mortality_F),pre_18_r_F,

mortality_F,dep_mortality_F,mortality_F,pre_18_d_F),

nrow = 4,

ncol = 4)

n = ncol(p)

A = t(p - diag(n))

A = rbind(A, rep(1, n))

b = c(rep(0, n), 1)

mu = qr.solve(A, b)

print(mu)

mu <- as.matrix(mu)

rownames(mu) <- c("healthy","depression", "remission", "death")

colnames(mu) <- c("prevalence")

#Recal Bias Female

lifetime_prevalence_F <- mu[2,1]+mu[3,1]

recall_bias_F <- (lifetime_prevalence_F/observed_lifetime_prevalence_F_low)-1

###Male

incidence_M_low=0.0013

incidence_M_high=0.0033

observed_lifetime_prevalence_M <- 0.151

p = matrix(c((1-mortality_M-incidence_M_low),0,0,pre_18_h_M,

incidence_M_low,(1-remission-dep_mortality_M),recurrence_M,pre_18_dep_M,

0,remission,(1-recurrence_M-mortality_M),pre_18_r_M,

mortality_M,dep_mortality_M,mortality_M,pre_18_d_M),

nrow = 4,

ncol = 4)

n = ncol(p)

A = t(p - diag(n))

A = rbind(A, rep(1, n))

b = c(rep(0, n), 1)

mu = qr.solve(A, b)

print(mu)

mu <- as.matrix(mu)

rownames(mu) <- c("healthy","depression", "remission", "death")

colnames(mu) <- c("prevalence")

#Recal Bias Male

lifetime_prevalence_M <- mu[2,1]+mu[3,1]

recall_bias_M <- (lifetime_prevalence_M/observed_lifetime_prevalence_M_low)-1
